# Computational simulation of virtual patients reduces dataset bias and improves machine learning-based detection of ARDS from noisy heterogeneous ICU datasets

**DOI:** 10.1101/2022.12.02.22283033

**Authors:** Konstantin Sharafutdinov, Sebastian Johannes Fritsch, Mina Iravani, Pejman Farhadi Ghalati, Sina Saffaran, Declan G. Bates, Jonathan G. Hardman, Richard Polzin, Hannah Mayer, Gernot Marx, Johannes Bickenbach, Andreas Schuppert

**Author notes:** CORRESPONDING AUTHOR: Konstantin Sharafutdinov.

## Abstract

**Goal:** Machine learning (ML) technologies that leverage large-scale patient data are promising tools predicting disease evolution in individual patients. However, the limited generalizability of ML models developed on single-center datasets, and their unproven performance in real-world settings, remain significant constraints to their widespread adoption in clinical practice. One approach to tackle this issue is to base learning on large multi-center datasets. However, such heterogeneous datasets can introduce further biases driven by data origin, as data structures and patient cohorts may differ between hospitals.

**Methods:** In this paper, we demonstrate how mechanistic virtual patient (VP) modeling can be used to capture specific features of patients’ states and dynamics, while reducing biases introduced by heterogeneous datasets. We show how VP modeling can be used to extract relevant medical information on individual patients with suspected acute respiratory distress syndrome (ARDS) from observational data of mixed origin. We compare the results of an unsupervised learning method (clustering) in two cases: where the learning is based on original patient data and on data ‘filtered’ through a VP model.

**Results:** More robust cluster configurations were observed in clustering using the VP model-based filtered data. VP model-based clustering also reduced biases introduced by the inclusion of data from different hospitals and was able to discover an additional cluster with significant ARDS enrichment.

**Conclusions:** Our results indicate that mechanistic VP modeling can be used as a filter to significantly reduce biases introduced by learning from heterogeneous datasets and to allow improved discovery of patient cohorts driven exclusively by medical conditions.

**IMPACT STATEMENT:** Mechanistic virtual patient modeling can be used as a filter to extract relevant medical information on individual patients, significantly reducing biases introduced by learning from heterogeneous datasets and allowing improved discovery of patient cohorts driven exclusively by medical conditions.

## I. Introduction

**A**rtificial intelligence (AI) and machine learning (ML) models have already shown their potential applicability in diverse areas of healthcare [1-3]. Several models have been developed for the early diagnosis and prediction of critical states and conditions in the ICU, e.g. ARDS [4], sepsis [5] and COVID-19 [6-9].

However, the more data-driven models are applied in healthcare settings, the more the issue of impaired performance on different datasets, i.e. poor generalizability of such models, is becoming apparent [5, 10-13]. If ML models are developed on one dataset, they learn data distributions which are specific or characteristic for this particular dataset, and perform worse on data obtained from other sources with potentially different distributions [14-16]. Moreover, attempts to apply models developed in a single hospital to patients from another hospital have also already revealed significant limitations [17, 18]. In medicine generally, but particularly in the ICU setting, there are multiple reasons why data from different hospitals can differ significantly, e.g. different admission strategies, guidelines for treatment, patients’ baseline values, protocols on settings of medical support devices or definitions of cut-off values [19-21].

On the one hand, the issue of poor generalizability of developed models cannot be solved by blindly increasing the size of the training dataset, as this does not necessarily guarantee a good performance of a model on another dataset [10]. On the other hand, pooling of data from diverse origins for development of AI/ML tools introduces further biases driven by data origin. This can represent a challenge for the application of both supervised and unsupervised AI/ML methods, as relevant medical information can be hidden behind biases introduced by different datasets [22].

A potential solution to these challenges is to exploit models that allow extraction of the core information describing a patient’s status. Such computer models, which are complex enough to model heterogeneous human pathophysiological states, are often referred to as “virtual patient (VP) models” or “in silico” patients [23]. These mechanistic models are based on well accepted and understood physiological principles and can be adapted to represent individual patients. They rely on real patient data and represent a specific pathophysiological state of a patient. Therefore, they can be considered a “digital twin” of a real patient at a given point in time [24]. VP models aim to capture specific features of patient dynamics while avoiding excessive detail. In other words, VP models can be conceptualized as “filters” to process heterogeneous data and extract information (parameters) which are essential to describe a patient’s state.

In this paper, we investigate how a mechanistic VP model can be employed as a model-based filter for ICU data pooled from diverse hospitals. We show that such model-based filtering allows a reduction in the bias introduced by diverse datasets, and provides clinically relevant information from noisy heterogeneous data, for instance from data pooled from different hospitals. We demonstrate our approach on a cohort of patients with suspected acute respiratory distress syndrome (ARDS) - a potentially life-threatening condition leading to respiratory insufficiency with relevantly impaired pulmonary gas exchange and possible multi-organ failure and fatal outcomes [25, 26] assessed from multiple hospitals in Germany as part of the ASIC project [27].

Despite the existence of an explicit clinical definition (the Berlin definition [28]), significant numbers of patients with ARDS are unrecognized or recognized late by clinicians [29-31]. Patients must fulfil a set of clinical criteria within a specific time frame that have relatively high sensitivity but low specificity, while the inter-observer reliability of the Berlin ARDS definition is moderate, mainly due to variability in chest X-ray interpretation. In addition, the oxygenation criterion, namely the ratio of arterial PO2 to inspired oxygen fraction, is not measured at standardized ventilator settings, and can vary substantially in a single patient for different FiO2 levels. Failure to recognize ARDS in a timely fashion leads to failure to use strategies that improve survival [31]. Early diagnosis of ARDS may facilitate measures to avoid progression of the lung injury, including protective mechanical ventilation, fluid restriction, and adjunctive measures proven to improve survival such as prone positioning.

Therefore, there is an urgent need for methods that could assist clinicians in early recognition of ARDS in the ICU setting. ML models for early recognition of ARDS developed to date have shown limited positive predictive value and have not been judged ready for clinical implementation [31]. In fact, under-recognition of ARDS by clinicians represents an important challenge to successful development of applicable ML models, since it causes insufficient quality of ARDS labeling in retrospective datasets, which in turn prevents the development of reliable ML models. In this paper we provide a way to address this issue. We show that clinically relevant information about individual patients can be extracted from raw data using a mechanistic VP model, and used to identify non-diagnosed ARDS patients, providing a route to improved ML model development for early ARDS recognition.

## II. Materials and Methods

### A. Computational model

The simulator used in this study includes a comprehensive simulation model of the pulmonary system based on mechanistic models of ventilation and gas exchange [32]. It was later extended to include cardiovascular components [33]. The simulator has already been validated using real patient data [34, 35]. Internally, the model is constructed as a system of differential algebraic equations obtained from published literature, experimental data, and observational studies, that quantitatively represent established physiological processes. The equations are solved iteratively, with the solutions of one iteration at a time point used as inputs to the iteration at the next time step. This allows accurate representation and observation of gradual changes in several parameters that are otherwise difficult to estimate. The simulator consists of different modules representing the airways, the lung as a collection of ventilated alveolar compartments coupled to mechanical ventilator, anatomical shunt, dead space and the tissue compartment. The lung is modeled using 100 alveolar compartments, each of which may have different properties such as flow resistance, vascular resistance, compliance, etc. Thus, ventilation-perfusion mismatch can be modeled, allowing the simulation of conditions such as ARDS [36-38].

The simulator represents a dynamic cardiopulmonary state in vivo that is initialized with numerous input parameters. Some of these parameters are routinely measured in intensive care setting, such as blood gas analysis (BGA) measurements or respirator settings (the full list of parameters used as inputs for the model is given in the Supplementary List I). Others, however, are rarely measured, such as cardiac output, anatomical shunt or biophysical characteristics of individual alveolar compartments, and thus these must be estimated using optimization procedures.

### B. Creation of a virtual patient cohort

To fully define each of the virtual patients, the simulator was fitted to individual patient data using advanced global optimization algorithm [39-41]. The model parameters that were identified in the optimization procedure included 2 groups of parameters. Firstly, rarely measured physiological parameters (anatomical shunt, respiratory quotient, anatomical dead space volume, metabolic rate of O2, cardiac stroke volume, and inspiration to expiration ratio), were determined through optimization if they were missing in patient data. Parameters defining distributions of properties of alveolar compartmental parameters (vascular resistance and flow resistance of compartments) were also identified in the optimization process. During the development of ARDS, due to an inflammatory process and a diffuse damage of alveolar-capillary membrane, protein-rich fluid enters the alveolar space impairing gas exchange. The weight of such a “wet lung” leads to an increased gravitational pressure on the lower, dependent lung compartments. This pressure in combination with the already present edema leads to the formation of atelectases, especially under mechanical ventilation (MV) with inadequate settings [42-44]. To model ARDS development, another main parameter was introduced to the optimization procedure – the number of closed alveolar compartments (n_cc_), accounting for the formation of atelectases and modeled through increased external pressure on the compartment leading to no ventilation and complete alveolar shunt. The optimization problem was formulated to find a configuration of model parameters that minimizes the difference between the model outputs and the observed patient data (arterial blood gas values at all time points in a window). Further details on the optimization procedure are given in the Supplementary File.

The optimization procedure was performed in two time windows relative to the onset of ARDS (t0): from t_0_ - 2d to t_0_ - 1d (window 1) and from t_0_ to t_0_ + 1d (window 2), where d stands for 1 day. The optimal parameterization of the simulator for each patient in the window 1 comprised a VP configuration. To model ARDS development, in the window 2 optimization was performed exclusively for the n_cc_ keeping the VP configuration found in the first window intact.

After fitting the simulator to individual patients, a list of parameters was calculated based on simulator outputs and parameters found in the optimization procedure in both time windows for each of the patients. These parameters, among others, included n_cc_, ventilation and shunted blood fraction (the full list of optimized and simulation output parameters is given in the Supplementary List II). For each of the patients, these parameters comprised model-based filtered data consisting of 18 features.

### C. Data

Four German hospitals (later referred to as Hosp A, Hosp C, Hosp D and Hosp E) provided retrospective, fully depersonalized data on ICU patients collected during the project “Algorithmic surveillance of ICU patients with acute respiratory distress syndrome” (ASIC) [27] of the SMITH consortium, which is part of the German Medical Informatics Initiative. The ASIC project was approved by the independent Ethics Committee (EC) at the RWTH Aachen Faculty of Medicine (local EC reference number: EK 102/19, date of approval: 26.03.2019). The ASIC project was registered at the German Clinical Trials Register (Registration Number: DRKS00014330). The Ethics Committee waived the need to obtain Informed consent for the collection and retrospective analysis of the de-identified data as well as the publication of the results of the analysis. Additionally, a historic retrospective dataset from one of the participating hospitals was included into the analysis (Hosp B). It comprised fully depersonalized data of ICU patients that were extracted according to the same rules as within the ASIC project. The time period for the historical dataset started with the introduction of the patient data management system in the ICU of the respective hospital and ended with the start of the ASIC project and covered a period of 10 years. Patient inclusion criteria were age above 18 years and a cumulative duration of invasive MV of at least 24 hours. There were no explicit exclusion criteria. Each patient’s data included routinely charted ICU parameters collected over the whole ICU stay, biometric data and ICD-10 codes. The full list of parameters used in this study is given in Supplementary List I. Data from all five datasets were brought to the same units of measurement and were checked for consistency. During depersonalization, the concept of k-anonymity was applied to several parameters that posed a risk to privacy. Due to this, not all datasets of patients who initially met the inclusion criteria could be extracted from the respective hospital and included in the final dataset. The overall number of patients in the final dataset comprised 29,275 patients.

The criteria for the diagnosis of ARDS are defined in the Berlin criteria [28]. As medical imaging data were missing in our dataset, only suspected ARDS onset time could be determined according to the Berlin criteria. It was defined as the timepoint when the ratio of arterial partial pressure of oxygen (PaO_2_) and the inspired fraction of oxygen (FiO_2_), also known as P/F ratio or Horovitz index, dropped below 300 mmHg for the first time and stayed below this threshold for at least 24 hours. Moreover, to be able to fit a simulator to the ICU data and create a cohort of virtual patients, only patients having specific MV, blood gas analysis and other parameters charted both before and after the suspected ARDS onset were selected. The final number of patients fulfilling these criteria comprised 1,007 patients. The number of patients before and after filtering in corresponding hospitals is given in Table I. A full description of data preparation and filtering is given in the Supplementary File.

**TABLE I.**
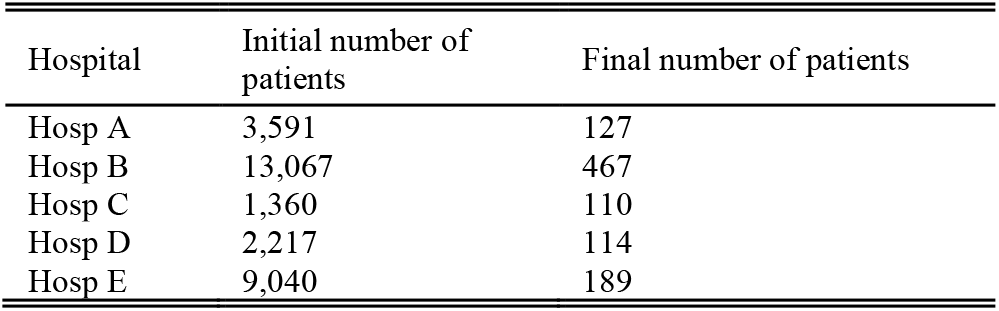
Initial and final number of patients in the hospitals under consideration.

| Hospital | Initial number of patients | Final number of patients |
| --- | --- | --- |
| Hosp A | 3,591 | 127 |
| Hosp B | 13,067 | 467 |
| Hosp C | 1,360 | 110 |
| Hosp D | 2,217 | 114 |
| Hosp E | 9,040 | 189 |

### D. Consensus clustering and enrichment analysis

We generated two datasets from the patient data representing the individual disease status to be used in the clustering algorithm. The first dataset comprised mean values of original measured parameters, which were used as inputs to the simulator, calculated on time windows 1 and 2 (before and after suspected ARDS onset respectively, see Supplementary List III). The second dataset comprised model-based filtered data: simulator outputs and parameters found in the optimization procedure (see Supplementary List II). The former dataset thus represented data from the cohort of original patients, while the latter represented the model-based filtered data or data from the virtual patient cohort.

Consensus k-means clustering was performed for different number of clusters in each of the cases. Consensus clustering is based on repeated multiple times (1000 times) clustering of the sampled data from the original dataset and is known to produce robust clusters [45]. To further increase robustness of discovered clusters, another step was introduced to the clustering procedure. It was allowed to assign an outlier label to some patients, if they could not be securely assigned to any of observed clusters. In the clustering procedure, quality of clustering was assessed using mean cluster’s consensus, as described in [45]. This metric is introduced based on consensus matrix D:

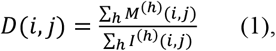

where M^(h)^ is a connectivity matrix of the perturbed dataset obtained in the h-th resampling of the original dataset and M^(h)^(i, j) is equal to 1, if items i and j belong to the same cluster in h-th clustering repetition and 0 otherwise. I^(h)^ is the (N × N) indicator matrix such that its (i, j)-th entry is equal to 1 if both items i and j are present in the perturbed dataset and 0 otherwise. Then, a cluster’s consensus m(k) is defined as the average consensus index between all pairs of items belonging to the same cluster k:

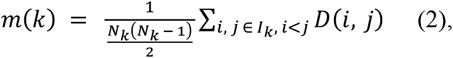

where I_k_ is the set of indices of items belonging to cluster k and N_k_ is a number of items in cluster k. Finally, the mean cluster’s consensus is the cluster’s consensus averaged over all clusters. This metric is a summary statistic which reflects the mean stability of clusters discovered in the consensus clustering algorithm and represents the overall robustness of discovered configuration of clusters. Mean clustering quality with 95 % confidence intervals was calculated by repeated (100 times) clustering on subsamples (80%) of dataset. A full description of the clustering procedure is given in the Supplementary File.

For each of the discovered clusters, enrichment with respect to clinical conditions and to underlying hospitals was evaluated using hypergeometric tests. Analogously to gene set enrichment analysis, this method allows to identify clinical conditions that are over-represented in a particular cohort (cluster) of patients compared to the whole population. Observed statistical significance values for each of conditions under consideration were corrected for multiple testing using Benjamini-Hochberg correction [46].

### E. Modules used in the study

In this study, the RBFOpt package [39] was used for fitting the VP model to real patient data in the optimization procedure. The following Python programming language [47] implementations were used in the study: scikit-learn [48] implementation of k-means clustering was used in the consensus clustering algorithm (sklearn.cluster.KMeans); scipy [49] implementations of hierarchical clustering were used in the consensus clustering algorithm (scipy.cluster.hierarchy, scipy.spatial.distance); statistical analysis was performed with scipy library (scipy.stats.hypergeom, scipy.stats.ttest_ind). Clustering results were compared using a two-tailed Student’s t-test with a significance level of α = 0.05.

## III. Results

### A. Optimization results

Fitting quality of the optimization procedure for all patients is shown in Fig. 1. Acceptable quality of fitting (simulator outputs within 2 standard deviations of measured data) was observed for 95.9% patients in the window before suspected ARDS onset and for 84.5% patients in the time window after suspected ARDS onset. Acceptable quality of fitting in both windows was observed for 81.7% or 823 patients, which were used in the subsequent analysis.

**Fig. 1.**
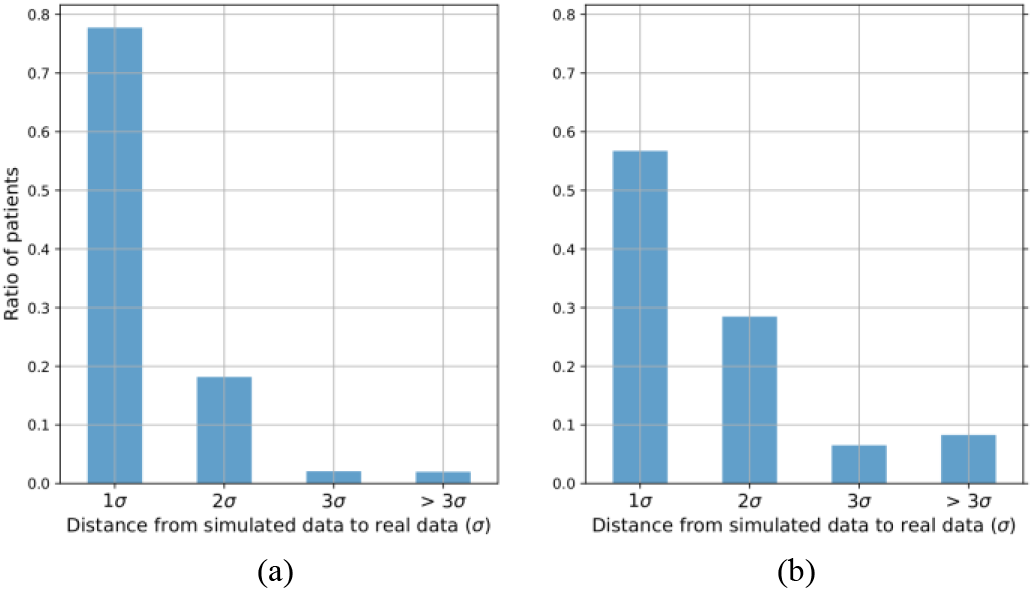
Quality of fitting the simulator to real patient in the time window before suspected ARDS onset (a) and after suspected ARDS onset (b). Cohort of 1007 patients with suspected ARDS. Acceptable quality of fitting (simulator outputs within 2 standard deviations of measured data) was observed for 95.9% patients in the window before suspected ARDS onset and for 84.5% patients in the time window after suspected ARDS onset.

### B. Clustering on original measured data

Clustering quality for different configurations of the number of clusters is shown in Fig. 2. The best clustering quality was observed for 2 clusters, followed by a steep decrease in clustering quality for 3 clusters and gradual decrease of clustering quality for clustering configurations with a cluster number larger than 5. Therefore, the number of clusters for further investigation was fixed to 5.

**Fig. 2.**
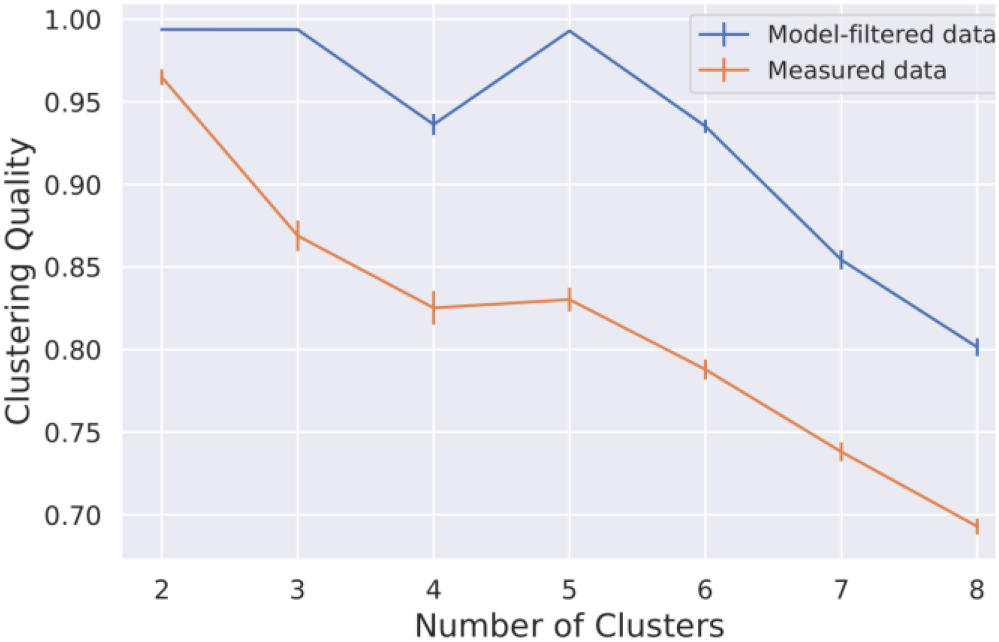
Clustering quality for different numbers of clusters for clustering on original measured data (orange line) and model-filtered data (blue line) data. Mean clustering quality with 95 % confidence intervals over repeated (100 times) clustering on subsample (80%) of dataset is shown. Mean clustering quality and results of a two-tailed Student’s t-test for mean quality of clustering are given in Table II.

Each of the 5 discovered clusters had characteristic clinical conditions, which were over-represented in the respective clusters. However, all clusters were found to be driven by data from one or several particular hospitals, i.e. significant enrichment with respect to the hospital was found. Furthermore, 4 out of 5 clusters were dominated by significant over-representation of underlying hospitals, i.e. the highest enrichment was observed with respect to the hospital and not to the medical condition, see Fig. 3 (a). Enrichment results are given in Supplementary Table I. Finally, none of the discovered clusters had significant enrichment of diagnosed ARDS patients (according to ICD-10 code J80.x).

**Fig. 2.**
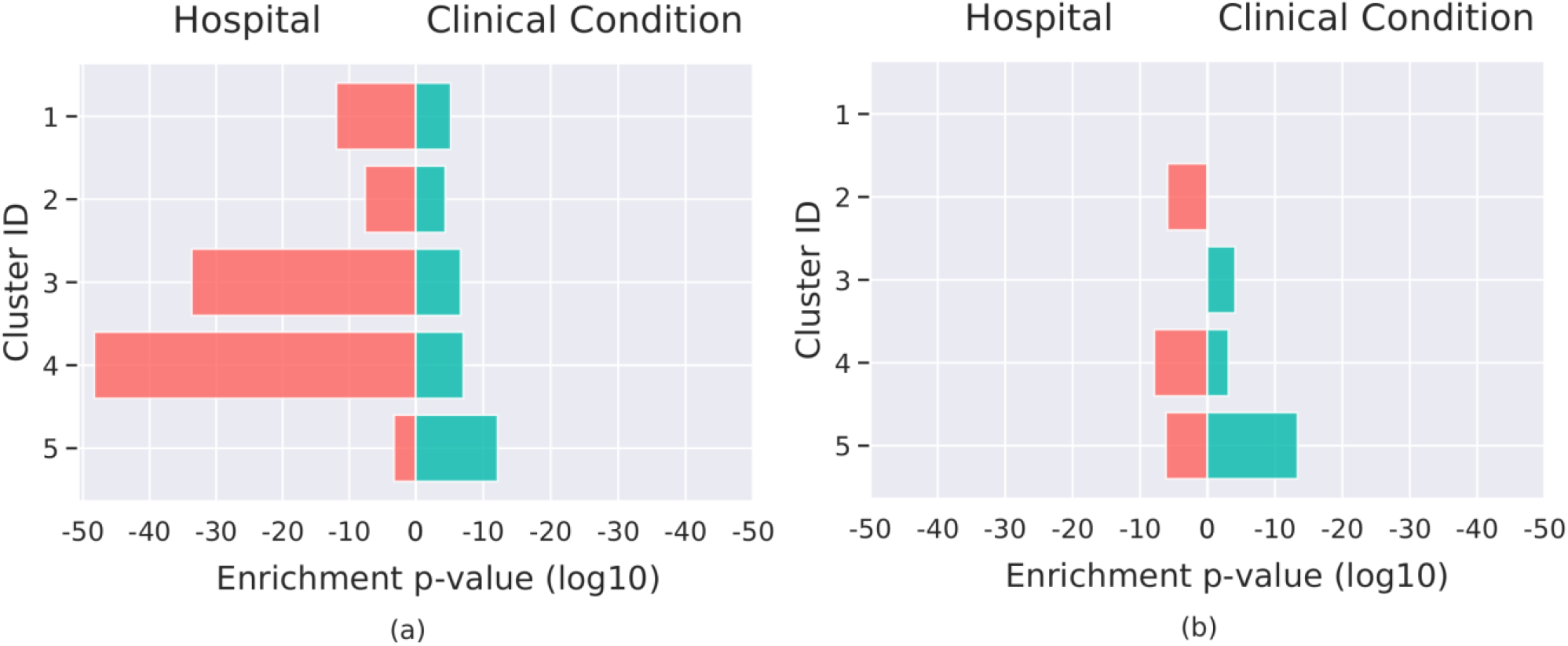
Significance of enriched clinical conditions and hospitals in discovered clusters for clustering on original measured data (a) and model-based filtered data (b). The highest enrichment in each of the clusters is shown both for enrichment of clinical conditions (green bar) and for enrichment with respect to hospital (red bar). In clustering on original data, all 5 discovered clusters are significantly enriched with data from some hospitals. In clustering on simulation data, 2 clusters without enrichment for a hospital are observed and overall magnitude of enrichment with respect to a hospital is decreased.

### C. Clustering on model-filtered data

In contrast to the clustering on the original measured data, the clustering quality on model-based filtered data was found to be significantly higher for all configurations of number of clusters (see Fig. 2 for the results of clustering, Table II for the results of the t-test, and Supplementary Table II for enrichment results). While on the original measured data, the quality decreased significantly already after increasing the number of clusters to 3, in the model-based filtered data, the quality remained high for 2, 3 and 5 clusters. However, a cluster number above 5 also resulted in a steep decrease in clustering quality in this dataset, and thus the number of clusters for further investigation was fixed to 5, similarly to the case of clustering on the original data.

**TABLE II.** Clustering quality for configurations with different number of clusters in case of clustering on original measured data and model-based filtered data. Mean clustering quality with 95 % confidence interval and results of a two-tailed Student’s t-test for mean quality of clustering are shown.

| Number of Clusters | Mean Quality Measured | Mean Quality Simulated | Statistic | p-value |
| --- | --- | --- | --- | --- |
| 2 | 0.965 (0.960, 0.970) | 0.994 (0.993, 0.995) | 11.726 | 9.481E-21 |
| 3 | 0.869 (0.860, 0.878) | 0.994 (0.993, 0.995) | 26.205 | 1.103E-46 |
| 4 | 0.825 (0.815, 0.835) | 0.936 (0.930, 0.942) | 18.137 | 2.373E-41 |
| 5 | 0.830 (0.823, 0.837) | 0.993 (0.992, 0.994) | 43.713 | 3.222E-67 |
| 6 | 0.788 (0.782, 0.794) | 0.935 (0.931, 0.939) | 39.515 | 2.879E-88 |
| 7 | 0.738 (0.732, 0.744) | 0.854 (0.848, 0.860) | 28.077 | 6.148E-71 |
| 8 | 0.693 (0.688, 0.698) | 0.801 (0.796, 0.806) | 29.223 | 2.781E-73 |

Clustering on model-based filtered data revealed 2 mixed clusters, i.e. clusters without over-representation of any underlying hospital. In the remaining 3 clusters, although such an over-representation could be observed, it was significantly lower than in the clustering on measured original data, see Fig. 3 (b) (significance of 5.0E-49, 2.2E-34, 1.2E-12, 2.5E-8, 5.8E-5 in measured data vs. 1.3E-8, 6.9E-7, 1.2E-6 in model-based filtered data).

Additionally, clustering on model-based data was able to discover a cluster with significant ARDS over-representation of diagnosed ARDS patients. This group of patients exhibited multiple properties which are specific for ARDS patients. These encompass the lowest Horovitz index among all clusters, the lowest number of ventilation-free days and the highest mortality. Finally, this cluster showed the largest increase in number of closed alveolar compartments (n_cc_) among all clusters.

## IV. Discussion

Data which are gathered in the ICU setting consist of global indices and parameters that reflect the state of the lung, such as BGA values or MV settings. However, these features in reality represent surrogate markers for the real pathophysiological state of the patient, leading to a significant simplification of clinical reality. In essence, ICU data are based on systematic monitoring of the enormous complexity of mechanisms accompanying the occurrence and progression of acute syndromes in individual patients. The development of complex syndromes is controlled not only by the core processes of disease progression (often molecular), but also by a large number of covariates arising from a diverse genetic background, lifestyle, exobiotic stress factors, and comorbidities. Another important factor is the large number of medical interventions in the context of intensive care, such as drug administration or MV. All these factors form highly complex feedback systems, in which the patient’s condition causes and influences the interventions to be performed, which in turn influence the patient’s condition. Such interventions can differ significantly among diverse hospitals introducing additional hospital bias to the datasets [50, 51]. Subsequently, relevant medical signals about a patient’s state are often disturbed by noise or are missing completely. For instance, the human lung has inhomogeneous characteristics such as structural asymmetries and regional variations in ventilation and perfusion that cannot be captured by standard diagnostic methods.

To be able to extract relevant patient information, approaches of systems medicine and computational physiology can be used. Systems medicine aims to describe, model, and simulate living, medically relevant systems using methods similar to those used for complex technical processes. The main goal of computational physiology as a part of systems medicine is the adequate description of these relationships in a computationally efficient manner and the development of models that consider unique properties of the living organisms in response to their environment [23, 52]. One of the pillars of computational physiology is VP modeling. The overall VP approach relies on the ability to determine parameters from data that are both patient-specific and time-varying, accounting for variability within and between patients. The ability of VP models, when appropriately adapted, to create a digital twin for a real patient also enables assessment of patient-specific parameters that are not readily measurable (e.g., vascular resistances, transpulmonary pressures, anatomic shunt, etc.). These unmeasurable parameters contain potentially important information about the patient’s health status, which cannot be extracted from routinely measured ICU data due to the previously mentioned reasons [24].

In this paper, we demonstrate how a VP modeling framework can be applied to large ICU patient cohorts pooled from different hospitals to reduce dataset bias and to extract medically relevant information. First, we show how a mechanistic VP model can be used to extract model-based filtered data of individual patients with suspected ARDS. Secondly, we show how these data can be further utilized to improve clustering quality and discover medically relevant patient subpopulations.

A comprehensive physiological model, that was used in this study was already validated against real patient data [34, 35]. However, in the current study, the simulator was firstly used to create a large (>1000 patients) cohort of virtual patients based on the retrospective observational data pooled from different hospitals. VP model fitting to real ICU patients showed a reasonable fitting quality. Acceptable fit in both time windows was observed for 81.7% of the patients in the cohort. The larger ratio of patients with acceptable quality of fitting in the first window can be explained by the fact that 11 parameters were optimized in the window 1, whereas only 1 parameter, namely n_cc_, was determined in the window 2. Therefore, reliable model-based filtered data were extracted for 823 patients.

To demonstrate the utility of the obtained model-based filtered data, we used a classic unsupervised learning approach, namely clustering. We compared the clustering on original data vs. clustering on extracted model-based filtered data. Intermediate clustering quality was observed in the clustering on original data, meaning that the consensus clustering method was struggling to split a full cohort into homogeneous groups and find a stable configuration of clusters. In contrast, clustering on model-based filtered data revealed significantly better clustering quality for all configurations of number of clusters.

More importantly, clustering based on the original data was strongly affected by the diversity of underlying hospitals. In all discovered clusters, patients from a particular hospital were significantly over-represented. In 4 out of 5 clusters, such enrichment was found to be the most significant for that cluster. These observations indicate that clustering on observed data is dominated more by the hospital source and much less by underlying medical conditions. Therefore, clustering on the pooled data is biased by the data source and does not allow to find mixed subgroups of patients. This finding is even more striking given the fact that we did not use external ICU datasets for this study, which could have covered different patient populations. All patients in this study satisfied the same strict inclusion criteria and were later filtered and chosen according to uniform rules. However, clustering on model-based filtered data obtained from each of the virtual patients allowed us to find 2 clusters of mixed hospital origin, i.e. clusters without over-representation of any underlying hospital. Moreover, although significant enrichment with respect to the hospital was still present in 3 out of 5 clusters, its magnitude was much less than in the clustering on original data (see Fig. 3).

These findings support the main characteristic of the VP models, namely the ability to identify relevant data patterns and extract hidden medical information from underlying data by leveraging mechanistic physiological principles while simultaneously avoiding an excessive level of detail.

Another interesting observation was that clustering on original measured data was not able to find a subgroup of “true” diagnosed ARDS patients. Partially, these patients were uniformly distributed among discovered clusters and did not form a separate group with typical ARDS properties, e.g. an impaired oxygenation or high driving pressures for MV. In contrast, clustering on model-based data was able to discover a cluster with significant ARDS over-representation and clinical properties, which resemble those of ARDS patients.

This finding is especially important in the context of unreliable ARDS labeling in retrospective data. Insufficient quality of labeling represents an additional factor that contributes to impaired generalization of AI/ML models developed on retrospective ICU data. For the proper development of ML models for ARDS diagnosis and prediction, such models have to be trained on reliably labeled data. On the one hand, patients labeled with ARDS ICD codes still represent a lower bound on the number of true ARDS cases, as large numbers of ARDS patients are not diagnosed [29-31]. On the other hand, reliable retrospective labeling constitutes a challenging task, due to the fact that diagnosis according to the Berlin definition requires the clinical appraisal of certain conditions, such as hypervolemia, which are not assessable retrospectively. Moreover, medical imaging data are frequently lacking in retrospective databases with observational ICU data. However, even if imaging data are available, reliable identification of the ARDS event remains a challenge due to a high interrater variability in chest imaging [53]. Finally, studies on the development of AI models for ARDS are utilizing diverging rules to retrospectively label ARDS patients [54-56].

All patients in the cohort under consideration had a time point (suspected ARDS onset), when a part of the Berlin definition which accounts for the impaired oxygenation was satisfied. Presence of “true” ARDS patients in the cohort was guaranteed by the fact, that some patients had ICD-10 code for diagnosed ARDS. However, some of the patients might have had ARDS, but were not diagnosed and therefore lacked the ICD-10 code for ARDS, since it is known that a relevant number of ARDS cases stays undiagnosed. Therefore, the “true” ARDS cohort would have consisted of these two groups of patients: the “true positives” and “false negatives”. Our hypothesis was that the patients from these two groups would be similar to each other and form a shared cluster in the clustering procedure. However, that was not the case for the clustering on original measured data, as none of the discovered clusters was enriched with diagnosed ARDS patients. Clustering on measured data was therefore not able to differentiate between ARDS patients and patients with other conditions, that could have led to decreased Horovitz index. In contrast, through clustering on model-based data we were able to discover a cluster with significant ARDS over-representation and clinical properties, which resemble those of ARDS patients. At the same time this cluster was not enriched with other pathological conditions, which often have similar clinical picture, such as for instance Heart Failure [57]. Furthermore, this ARDS cluster had the largest increase in the number of closed compartments (n_cc_) in the model, which fully supports our approach of modeling ARDS by introducing closed alveolar compartments. Our findings suggest that the identified ARDS cluster might also include those ARDS patients which were not diagnosed by the ICU staff. Therefore, this model-based filtering approach could be additionally used to identify non-diagnosed ARDS patients, although further research and retrospective validation is needed to prove this hypothesis.

Our study has some limitations that have to be considered. Parameters of the virtual patients that were identified in the window before suspected ARDS onset were assumed to stay constant in the observation window of 2 days. This is only partially true, as most of the identified parameters are changing with time. Therefore, our approach to model ARDS development represents a significant simplification of the complex pathophysiological processes, which are happening during this critical condition. However, in our opinion, it covers the most important clinical manifestation of ARDS and can be used as the first approximation for the modeling. Moreover, our ARDS modeling approach was validated by the fact that the ARDS cluster, which was discovered in the data, had the largest increase in number of closed compartments, as expected. Nevertheless, VP modeling has the potential to extract a lot of additional information about the patient status which was not used in this study. By introducing physiologically meaningful changes in other VP parameters during ARDS development, one might significantly improve quality of ARDS modeling.

Extensive data requirements and complexity of the fitting process of the VP model constituted additional limitations of the study. The former did not allow us to use all available patient data and was the reason for the significantly lower of number of patients in the final analysis cohort compared to the initial cohort (see Table I). It must be considered that to reach the aim to create a sufficiently large dataset for the analysis, not only data collected during the current project but also older datasets were included. It cannot be ruled out that patient populations or therapeutic concepts have changed over the years introducing additional bias into the analysis. However, this limitation reflects the real-world situation, as ML models are mostly developed on retrospective datasets with some temporal separation from datasets, where such models are intended to be used. Furthermore, this limitation does not influence the overall conclusions of the study, as enrichment of a similar magnitude was observed with respect to this dataset and to 4 datasets from other hospitals. The latter limitation required the use of the computing cluster for the optimization procedure. Although our approach was limited only to the identification of at most 11 parameters for each of the virtual patients, it required the use of advanced global optimization algorithm and significant computational resources. All this still tremendously complicates a straightforward implementation of such methods at the bedside.

In general, VP modeling possesses further limitations, restraining its applicability in real-world setting. First, it requires complex validation of the developed models [24]. Second, VP models are usually limited to an organizational level of the human body and do not consider the influence of exogenous covariates, e.g., preexisting diseases, lifestyle, genetic predispositions, or environmental influences. This conceptual problem can be solved using a hybrid modeling approach [58, 59].

## V. Conclusions

In this study we have shown how a mechanistic VP model can be used to extract relevant medical information on individual patients with suspected ARDS from observational data of mixed origin. Our results support the hypothesis that mechanistic modeling can be used as a filter to significantly reduce biases in data, introduced by pooling of data from different hospitals and to allow a discovery of patient cohorts driven exclusively by medical conditions. Overall, the continuous development of hybrid modeling approaches integrating diverse computational technologies, continuing increases in computational power, and ever-growing numbers of available datasets leads to the expectation that these technologies will make a significant contribution to precision medicine, with benefits for patients, physicians, and the healthcare system as a whole.

## Data Availability

All data produced in the present study are available upon reasonable request to the authors.

## Supplementary Materials

Supplementary materials include description of data preparation and filtering, optimization, and clustering approaches used in the study. Supplementary List I contains the full list of parameters used as inputs for the model. Supplementary List II contains the full list of optimized and simulation output parameters which comprise model-based filtered data. Supplementary List III contains the full list of features which were extracted from original measured data and used in the clustering procedure. Enrichment analysis results for each of discovered clusters in case of clustering on original measured data are given in Supplementary Table I. Finally, enrichment analysis results for each of discovered clusters in case of clustering on model-based filtered are given in Supplementary Table II.

## Acknowledgment

This publication of the SMITH consortium was supported by the German Federal Ministry of Education and Research (Grant Nos. 01ZZ1803B and 01ZZ1803M).

## Author contributions

JGH, SS and DGB developed the VP ARDS model. HM, SJF, KS, and RP worked on data acquisition and harmonization. KS and MI developed and implemented VP ARDS modeling pipeline. HM gave advice during development of the VP ARDS modeling framework. KS and PFG developed and implemented clustering routines. KS and AS designed the research and performed analysis of the patient data. SJF gave medical advice during the development of the pipeline. SJF, GM and JB interpreted the results from a medical perspective. KS, SJF, and AS wrote the manuscript. All authors read and approved the final manuscript.

