## Supplementary Material for "Computational simulation of virtual patients reduces dataset bias and improves machine learning-based detection of ARDS from noisy heterogeneous ICU datasets"

**Supplementary Materials**

Computational simulation of virtual patients reduces dataset bias and improves machine learning-based detection of ARDS from noisy heterogeneous ICU datasets

### I. Data preparation and filtering

Original patient cohort was filtered based on several criteria. First, patients with suspected ARDS were chosen. Suspected ARDS was defined when patient had a timepoint when the Horovitz index (PaO_2_/FiO_2_) dropped below 300 and stayed below this threshold for at least 24 hours. Second, only patient datasets with following parameters charted in both time windows before and after ARDS onset were chosen: PaO_2_, PaCO_2_, pHa, HCO_3_, FiO_2_, PEEP, P_EI_. Other parameters which are given in the Supplementary List I were also extracted from real patient data. If some of them were missing, they were substituted with median values for that parameter of the whole patient population. As SV and I:E were missing completely or in majority of patients in datasets from some of the hospitals, the values for these 2 parameters were identified in the optimization procedure if they were missing, as described above.

### II. Optimization procedure

The Nottingham Physiology Simulator (NPS V1.4) was used as a mechanistic virtual patient (VP) model in this study. This model and its components have been extensively described elsewhere [1]. Prior to the optimization procedure, patient data were binned into 2h bins. Then, patient datasets with charted values of the following parameters:

- PaO_2_
- PaCO_2_
- pH
- HCO_3_
- Fraction of inspired oxygen (FiO_2_)
- Positive end-expiratory pressure (PEEP)
- End-inspiratory pressure (P_EI_)

on at least 3 time points in each of the time windows were selected. To fully define each of the virtual patients, the simulator was fitted to individual patient data. Optimization task was formulated as minimization of the objective function Y with respect to parameters that have to be identified in the optimization procedure:

$\min_{p_{1},\ldots,p_{j}} \left( Y \right) =\min_{p_{1},\ldots,p_{j}} \left( \frac{\sum_{time points} \sqrt{\sum_{i = 1}^{3} r_{i}^{2}}}{n_{time points}} \right)$, $r_{i} = \frac{y_{i}^{meas} - y_{i}^{sim}}{\sigma_{i}}$	(1),

where y are arterial blood gas analysis parameters y = [PaO_2_, PaCO_2_, HCO_3_] and p_i_ are optimization parameters. Here, the difference between original measured values (y^meas^) and simulator outputs (y^sim^) was taken and scaled with the standard deviation of the corresponding parameter of a patient. Then, this difference was summed over all blood gas analysis parameters and all time points in the time window under consideration.

Parameters which were identified in the optimization procedure included:

- rarely measured physiological parameters (if missing in patient data):
- anatomical shunt (anatShunt)
- respiratory quotient (RQ)
- anatomical deadspace volume (VDphys)
- metabolic rate of O2 (VO2)
- stroke volume (SV)
- inspiration : expiration ratio (I:E)
- parameters defining distributions of properties of alveolar compartmental parameters:
- Distribution of vascular resistance in the lung was introduced through two parameters (sVR and inVR) defining values VR_i_ of vascular resistance of a compartment *i* for each of 100 alveolar compartments in the model based on VR values of a healthy patient: VR_i_ = VR_healthy_ + (i-49)*sVR + inVR
- Analogously, distribution of flow resistance in the lung was introduced through two parameters (sR and inR) defining values R_i_ of flow resistance of a compartment *i* for each of 100 alveolar compartments in the model based on R values of a healthy patient: R_100-i_ = R_healthy_ + (i-49)*sR + inR
- number of closed alveolar compartments (n_cc_). Closed compartments were introduced through setting values of external pressure of n_cc_ random compartments to large negative number (-30 cmH_2_O). Large negative external pressure models a scenario where there is compression from outside the alveolus, for instance through interstitial edema, causing collapse. Additionally, stiffness of alveoli of closed compartments was set to large value (0.16 cmH_2_O*ml^-2^), modeling stiffer lung units.

The optimization procedure was performed in two time windows relative to the onset of ARDS (t_0_): from t_0_ - 2d to t_0_ - 1d (window 1) and from t_0_ to t_0_ +1d (window 2), where d stands for 1 day. The optimal parameterization of the simulator for each patient in the window 1 comprised a VP configuration. Overall, maximum 11 parameters were optimized to completely define a virtual patient for a single real patient dataset. To model ARDS development, in the window 2 optimization was performed exclusively for the n_cc_ keeping the VP configuration found in the first window intact. The RBFOpt package was used as an optimizer in the study [2].

### III. Clustering

Clustering was performed on two different datasets. The first dataset comprised original data, which were used as inputs to the simulator (please see Supplementary List III for the list of features). For each of the parameters, mean values in windows 1 and 2 were calculated and used as features. Additionally, the mean Alveolar–arterial gradient (A-aO_2_ or A–a gradient) in each of the time windows was calculated and added to the list of features. Increased A–a gradient representing the large difference between the alveolar and arterial concentrations of oxygen implies ventilation-perfusion mismatch, therefore reflecting impaired integrity of the alveolar capillary unit. In states of ventilation-perfusion mismatch, such as pulmonary embolism or ARDS, oxygen is not effectively transferred from the alveoli to the blood which results in an elevated A-a gradient [3]. Finally, difference in Horovitz index between window 1 and window 2 was included to the list of features and used as a feature reflecting the severity of development of oxygenation impairment. Next, highly correlated features with a Pearson correlation coefficient between corresponding measurements larger than 0.9 were omitted from the analysis. The second dataset comprised model-based filtered data: simulator outputs and parameters found in the optimization procedure (please see Supplementary List II).

Consensus k-means clustering was performed for different number of clusters (from 2 to 8 clusters) in each of the cases. Consensus clustering is based on repeated multiple times (1000 times) clustering of the sampled data from the original dataset. Based on produced clusters a consensus matrix is formed, which reflects how many times two items of the dataset occurred in the same cluster. Then, hierarchical clustering is performed on the consensus matrix to produce final clusters. To further increase robustness of discovered clusters, another step was introduced to the clustering procedure. It was allowed to assign an outlier label to some patients, if they could not be securely assigned to any of the observed clusters based on the predefined threshold. The threshold for the final clustering configuration was chosen in the way, that at most 20% of patients were labeled as outliers.

In the clustering procedure, quality of clustering was assessed using mean cluster’s consensus, as described in [4]. This metric is a summary statistic which reflects the mean stability of clusters discovered in the consensus clustering algorithm and represents the overall robustness of discovered configuration of clusters. Mean clustering quality with 95 % confidence intervals was calculated by repeated (100 times) clustering on subsamples (80%) of dataset.

Clinical conditions which were used in the enrichment analysis were defined based on ICD-10 codes of underlying patients. Following clinical conditions were extracted from the codes: all ICD-10 chapters (A00-B99: Certain infectious and parasitic diseases, C00-D48: Neoplasms, etc..), all ICD-10 blocks (A00-A09: Intestinal infectious diseases, A15-A19 Tuberculosis, etc..) and comorbidities related to ARDS: Aspiration, Burn Trauma, Chronic Heart Failure, Chronic Renal Failure, Diabetes mellitus, Drug overdose, Hematologic Neoplasm, Immunosuppression, Inhalation, Liver Failure, Pankreatitis, Pneumonia, Renal Failure, Sepsis, TRALI, and Thoraxtrauma. Only clinical conditions with prevalence of more than 1% in the underlying population were used in the analysis.

Supplementary list I

Diagnostic parameters used in this study. Overall, 17 diagnostic parameters routinely assessed in the ICU were used in this study.

Vital signs:

- Body temperature

- Heart rate

- Stroke volume

Ventilatory settings:

- Fraction of inspired oxygen (FiO_2_)

- Inspiration : Expiration ratio (I:E)

- Positive end-expiratory pressure (PEEP)

- Tidal volume

- End-inspiratory pressure (P_EI_)

- Respiratory rate

Blood gas analysis parameters:

- PaCO_2_

- PaO_2_

- SaO_2_

- pH arterial (pHa)

- Bicarbonate (arterial) (HCO_3_)

- Base excess (arterial)

- PaO_2_/FiO_2_ ratio (Horovitz index)

Laboratory parameters:

- Haemoglobin

Supplementary list II

Parameters used as model-based filtered data. These were calculated based on simulator outputs or found in the optimization procedure in both time windows for each of the patients. For each of the patients these parameters comprised model-based filtered data consisting of 18 features. Values of these parameters were used as features in the clustering procedure.

- anatomical shunt (anatShunt)

- respiratory quotient (RQ)

- anatomical deadspace volume (VDphys)

- metabolic rate of O2 (VO2)

- slope of vascular resistance (sVR)

- intercept of vascular resistance (inVR)

- slope of flow resistance (sR)

- intercept of flow resistance (inR)

- number of closed compartments in window 1 (n_cc1_)

- number of closed compartments in window 2 (n_cc2_)

- fitting residual in window 1

- fitting residual in window 2

- lung ventilation in window 1

- ratio of shunted blood in window 1

- lung ventilation in window 2

- ratio of shunted blood in window 2

- increase in number of closed compartments (n_cc2_ - n_cc1_)

- volume of shunted blood through closed compartments in window 2

Supplementary list III

Features extracted from original measured data. Mean values of the following parameters in windows 1 and 2 (before and after suspected ARDS onset respectively) were calculated and used as features in the clustering procedure. Additionally, difference in Horovitz index between mean values in window 1 and window 2 was used as a feature (Horovitz drop). Overall, 31 features extracted from original measured data.

Vital signs:

- Body temperature

Ventilatory settings:

- Fraction of inspired oxygen (FiO_2_)

- Positive end-expiratory pressure (PEEP)

- Tidal volume

- End-inspiratory pressure (P_EI_)

- Respiratory rate

Blood gas analysis parameters:

- PaCO_2_

- PaO_2_

- SaO_2_

- pH arterial (pHa)

- Bicarbonate (arterial) (HCO_3_)

- Base excess (arterial)

- PaO_2_/FiO_2_ ratio (Horovitz index)

Laboratory parameters:

- Haemoglobin

Calculated parameters:

- Alveolar-arterial gradient

- Horovitz drop

Supplementary table I

Hospitals and clinical conditions which are over-represented in the discovered clusters. Results for clustering on original measured data are shown.

| Cluster ID | Condition/Hospital | p-value | Prevalence Cluster | Prevalence Population |
| --- | --- | --- | --- | --- |
| 1 | Hosp B | 1.164E-12 | 0.746 | 0.464 |
| 1 | Other disorders of the nervous system | 6.961E-06 | 0.500 | 0.328 |
| 1 | Cerebrovascular diseases | 5.595E-04 | 0.523 | 0.390 |
| 1 | Other disorders of eye and adnexa | 1.212E-03 | 0.138 | 0.068 |
| 2 | Hosp B | 2.529E-08 | 0.679 | 0.464 |
| 2 | Persons encountering health services for specific procedures and health care | 4.250E-05 | 0.723 | 0.570 |
| 2 | Hosp A | 5.811E-05 | 0.234 | 0.124 |
| 3 | Hosp D | 2.190E-34 | 0.466 | 0.123 |
| 3 | Hosp C | 3.234E-12 | 0.291 | 0.109 |
| 3 | Occupied and unoccupied key numbers | 2.223E-07 | 0.500 | 0.317 |
| 3 | Functional impairment | 3.545E-07 | 0.291 | 0.147 |
| 3 | Persons encountering health services for examination and investigation | 2.650E-05 | 0.514 | 0.363 |
| 3 | Codes for special purposes | 5.548E-05 | 0.878 | 0.758 |
| 3 | Injuries to the shoulder and upper arm | 3.947E-04 | 0.088 | 0.034 |
| 3 | Obesity and other hyperalimentation | 1.275E-03 | 0.149 | 0.080 |
| 3 | Thoraxtrauma | 2.002E-03 | 0.189 | 0.114 |
| 3 | Injuries to the thorax | 2.002E-03 | 0.189 | 0.114 |
| 3 | Injuries to the head | 2.940E-03 | 0.243 | 0.162 |
| 4 | Hosp E | 5.043E-49 | 0.636 | 0.180 |
| 4 | Persons encountering health services for examination and investigation | 8.975E-08 | 0.552 | 0.363 |
| 4 | Occupied and unoccupied key numbers | 1.033E-05 | 0.468 | 0.317 |
| 4 | Other diseases of intestines | 3.647E-05 | 0.318 | 0.196 |
| 4 | Chronic lower respiratory diseases | 7.238E-05 | 0.201 | 0.107 |
| 5 | Renal failure | 7.680E-13 | 0.792 | 0.454 |
| 5 | Aplastic and other anaemias | 1.696E-11 | 0.958 | 0.697 |
| 5 | Diseases of liver | 3.956E-09 | 0.396 | 0.164 |
| 5 | Liver Failure | 5.741E-09 | 0.333 | 0.124 |
| 5 | Diseases of the genitourinary system | 2.450E-08 | 0.865 | 0.621 |
| 5 | Diseases of the blood and blood-forming organs | 2.526E-08 | 0.969 | 0.775 |
| 5 | Sepsis | 4.334E-08 | 0.802 | 0.550 |
| 5 | General symptoms and signs | 5.499E-08 | 0.917 | 0.699 |
| 5 | Diseases of the digestive system | 9.884E-07 | 0.698 | 0.467 |
| 5 | Coagulation defects, purpura and other haemorrhagic conditions | 3.379E-06 | 0.760 | 0.546 |
| 5 | Other bacterial diseases | 4.122E-06 | 0.740 | 0.525 |
| 5 | Other diseases of the digestive system | 1.149E-05 | 0.250 | 0.106 |
| 5 | Mycoses | 7.332E-05 | 0.354 | 0.196 |
| 5 | Complications of surgical and medical care, not elsewhere classified | 8.169E-05 | 0.552 | 0.369 |
| 5 | Suppurative and necrotic conditions of lower respiratory tract | 3.792E-04 | 0.094 | 0.027 |
| 5 | Hosp B | 5.660E-04 | 0.625 | 0.464 |
| 5 | Diseases of oesophagus, stomach and duodenum | 8.531E-04 | 0.281 | 0.159 |
| 5 | Soft tissue disorders | 1.449E-03 | 0.156 | 0.070 |
| 5 | Diseases of the skin and subcutaneous tissue | 1.532E-03 | 0.438 | 0.299 |
| 5 | Diseases of peritoneum | 1.996E-03 | 0.188 | 0.095 |
| 5 | Other respiratory diseases principally affecting the interstitium | 2.685E-03 | 0.229 | 0.129 |
| 5 | Certain infectious and parasitic diseases | 4.320E-03 | 0.885 | 0.781 |
| 5 | Other disorders of the skin and subcutaneous tissue | 4.929E-03 | 0.365 | 0.249 |
| 5 | Symptoms, signs and abnormal clinical and laboratory findings, not elsewhere classified | 5.172E-03 | 0.948 | 0.865 |
| 5 | Infectious agents with resistance to certain antibiotics or chemotherapeutic agents | 6.836E-03 | 0.333 | 0.226 |
| 5 | Glomerular diseases | 7.804E-03 | 0.094 | 0.039 |

Supplementary table II

Hospitals and clinical conditions which are over-represented in the discovered clusters. Results for clustering on model-based filtered data shown. In cluster 1 no significant enrichment was found.

| Cluster ID | Condition/Hospital | p-value | Prevalence Cluster | Prevalence Population |
| --- | --- | --- | --- | --- |
| 1 | - | - | - | - |
| 2 | Hosp B | 1.212E-06 | 0.632 | 0.464 |
| 3 | Other respiratory diseases principally affecting the interstitium | 8.030E-05 | 0.269 | 0.129 |
| 3 | ARDS | 1.549E-04 | 0.247 | 0.118 |
| 4 | Hosp B | 1.308E-08 | 0.721 | 0.464 |
| 4 | Other bacterial diseases | 7.838E-04 | 0.673 | 0.525 |
| 4 | Aplastic and other anaemias | 1.040E-03 | 0.827 | 0.697 |
| 4 | Sepsis | 1.165E-03 | 0.692 | 0.550 |
| 5 | Occupied and unoccupied key numbers | 4.514E-14 | 0.640 | 0.317 |
| 5 | Persons encountering health services for examination and investigation | 1.129E-11 | 0.658 | 0.363 |
| 5 | Hosp E | 6.872E-07 | 0.360 | 0.180 |
| 5 | Codes for special purposes | 1.235E-05 | 0.910 | 0.758 |
| 5 | Hosp A | 4.379E-05 | 0.252 | 0.124 |
